# Predicting the end-stage of the COVID-19 epidemic in Brazil

**DOI:** 10.1101/2020.05.28.20116103

**Authors:** W.E. Fitzgibbon, J.J. Morgan, G.F. Webb, Y. Wu

**Affiliations:** Department of Mathematics, University of Houston; Department of Mathematics, Vanderbilt University; Department of Mathematics, Middle Tennessee State University

**Keywords:** asymptomatic transmission, symptomatic transmission, turning point, final size, COVID-19 epidemic in Brazil

## Abstract

We develop a dynamic model of a COVID-19 epidemic as a system of differential equations. The model incorporates an asymptomatic infectious stage and a symptomatic infectious stage. We apply the model to the current COVID-19 epidemic in Brazil. We compare the model output to current epidemic data, and project forward in time possible end-stages of the epidemic in Brazil. The model emphasizes the importance of reducing asymptomatic infections in controlling the epidemic.

## 1 Model

The epidemic population is divided into three classes: a susceptible class *S*, and two infected classes *I*_1_ and *I*_2_, representing the asymptomatic (or low level symptomatic) infectious individuals, and symptomatic (or high level symptomatic) infectious individuals, respectively. Susceptible individuals become asymptomatically infected via contact with either asymptomatically infectious or symptomatically infectious individuals. The transmission process is modelled by the force of infection term *S* (*τ*_1_*I*_1_ + *τ*_2_*I*_2_), which constitutes a loss rate for the susceptible class and a gain rate for the asymptomatic infected class. Asymptomatic individuals become symptomatic at rate λ. Symptomatic individuals are removed at rate γ, due to recovery, isolation, mortality, or other reasons. These elements lead to the following system of ordinary differential equations:

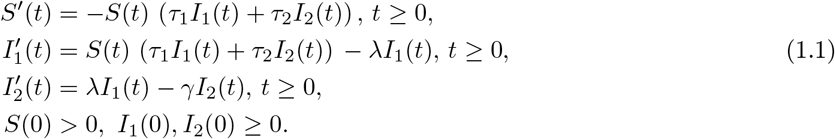

A flow diagram of the model is given in Figure 1. We remark that a model similar to (1.1) is developed in [12, 13, 14, 15], where *I*_2_ is further differentiated into reported and unreported classes. The global dynamics of (1.1) are given in the following theorem:

**Figure 1:**
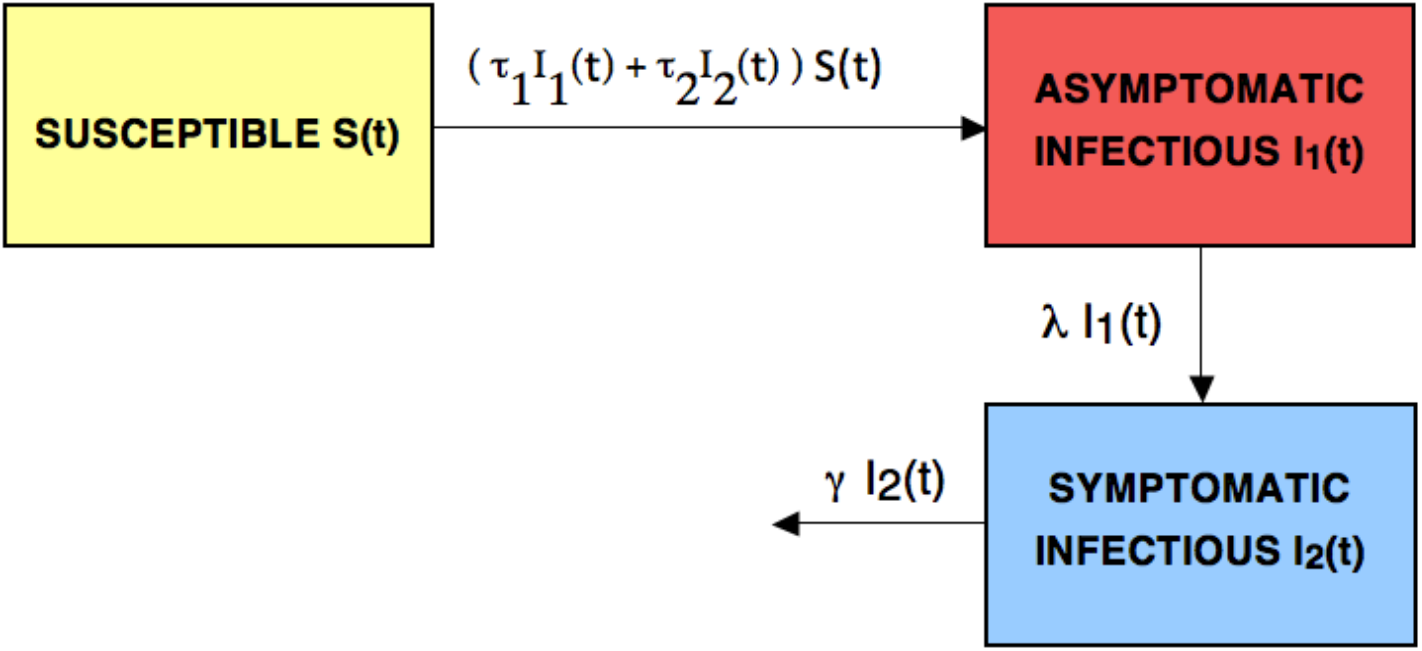
Flow diagram of the model.

### Theorem 1.1

*Suppose τ*_1_ > 0,*τ*_2_ > 0, λ > 0, *and* γ > 0. *Then* (*1.1*) has bounded nonnegative solutions *S*(*t*), *I*_1_(*t*), *I*_2_(*t*) *for t* ≥ 0, *and*

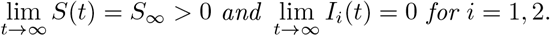

This result is proved in greater generality in [11], in a setting with *S, I*_1_ and *I*_2_ spatially dependent, and their equations replaced by reaction-diffusion equations over a geographical domain. For the spatially homogeneous case, we can express *S_∞_* and the basic reproductive number 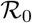 in terms of the initial conditions and parameters in the model (1.1):

#### Corollary 1.2

*S*_∞_ *satisfies the equation*

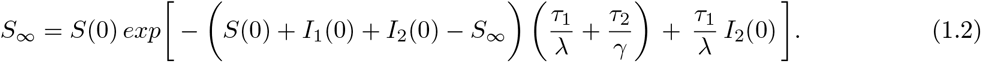

*The basic reproductive number is*

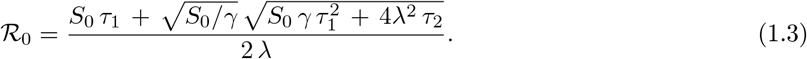

Proof.

From the first equation in (1.1) we obtain

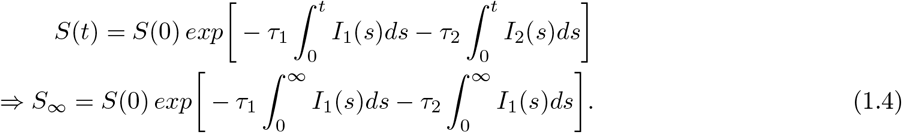

From the sum of the first, second, and third equation in (1.1) we obtain

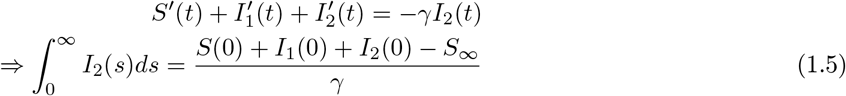

From the third equation in (1.1) we obtain

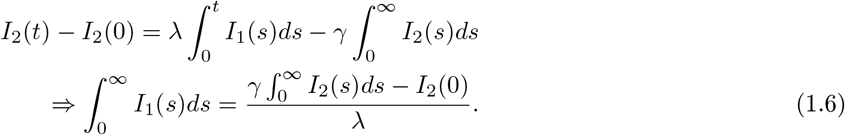

Then, (1.2) follows from (1.4), (1.5), and (1.6).

To prove (1.3) we use the next generation method [9, 10]. The linearized equations of the infectious part of the system are

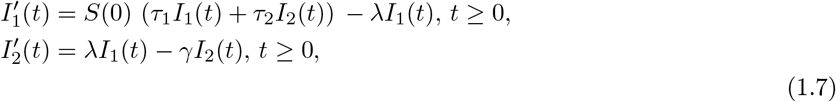

The corresponding matrix is

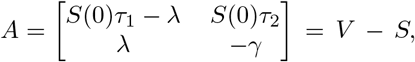

where

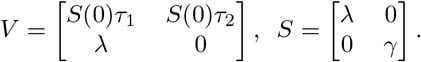

The next generation matrix is

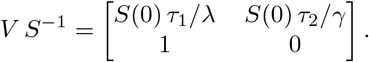

Then, 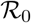 in (1.3) is the dominant eigenvalue of *V S*^−1^. ■

## 2 COVID-19 in Brazil

We apply this result to the COVID-19 epidemic in Brazil. Other models of the COVID-19 epidemic in Brazil are in [1, 2, 3, 4, 5, 6, 7, 8, 16, 17, 18, 19, 20]. Currently, the epidemic in Brazil is in a rapid growth phase, with limited social distancing measures in effect, and limited compliance with these measures. We use the cumulative daily reported cases data for this epidemic from the Ministerio da Saude of Brazil (https://coronavirus.saude.gov.br/).

We set the time units to days. We view *I*_1_(*t*) as the population of asymptomatic or low level symptomatic infectious individuals at time *t*. We view *I*_2_(*t*) as the population of high level symptomatic infectious individuals at time *t*. We set *S*(0) = 210,000,000, the current population of Brazil. We set the loss rate λ of *I_1_*(*t*) to 1/7 per day, which means *I*_1_ infectiousness lasts 7 days on average. We set the loss rate γ of *I*_2_ (*t*) to 1/7 per day, which means *I*_2_ infectiousness lasts 7 days on average. We believe the values λ = 7 and γ = 7 are reasonable estimates at this time for the current COVID-19 epidemic in Brazil. We identify an interval of time [*t*_0_,*t*_1_] on which the cumulative daily reported cases data is growing: *t*_0_ = 1, corresponding to March 17 and *t*_1_ = 75, corresponding to May 30.

The cumulative number *I*_1_(*t*) of low level infectious cases as a function of time *t* is 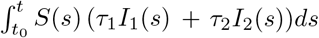. The cumulative number *I*_2_(*t*) of high level infectious cases as a function of time *t* is 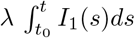. The high level infectious cases *I*_2_ are removed at the rate γ per day. We assume that a fraction *f* = 0.3 of these removed cases are reported, isolated, and cause no further transmissions. Thus, the cumulative number of reported cases at time *t* is 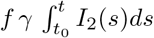, with *f* = 0.3 and γ = 1/7. Other fractional values *f* could be assumed, but currently this fractional value is not known.

We fit the parameters *τ*_1_, *τ*_2_, and initial values *I*_1_ (*t*_0_), *I*_2_(*t*_0_), so that the solutions of (1.1) align with the cumulative reported cases data on the time interval [*t*_0_,*t*_1_]. For this time interval, we estimate *τ*_1_ = 2.5 × 10^−10^, *τ*_2_ = 1.0 × 10^−9^, *I*_1_(*t*_0_) = 13,000, *I*_2_(*t*_0_) = 8, 200, by fitting the solutions of (1.1) to the cumulative reported cases data between March 17 and May 30. This means that 25% of transmissions are due to *I*_1_ asymptomatic or low level symptomatic cases, in this stage of the epidemic, in which the number of daily reported cases in increasing. In Figure 2 we graph the cumulative reported cases data and the cumulative reported cases from the simulation of the model. Other choices of the parameters and initial values are possible.

The daily reported cases from the model simulation, with *f* = 0.3, γ = 1/7, *t*_0_ = 1 (March 17), is obtained by solving the differential equation

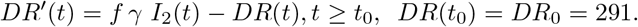

In Figure 3 we graph the daily reported cases data and the daily reported cases from the model simulation with *f* = 0.3, γ =1/7 and *t*_0_ = 1 (March 17).

**Figure 2:**
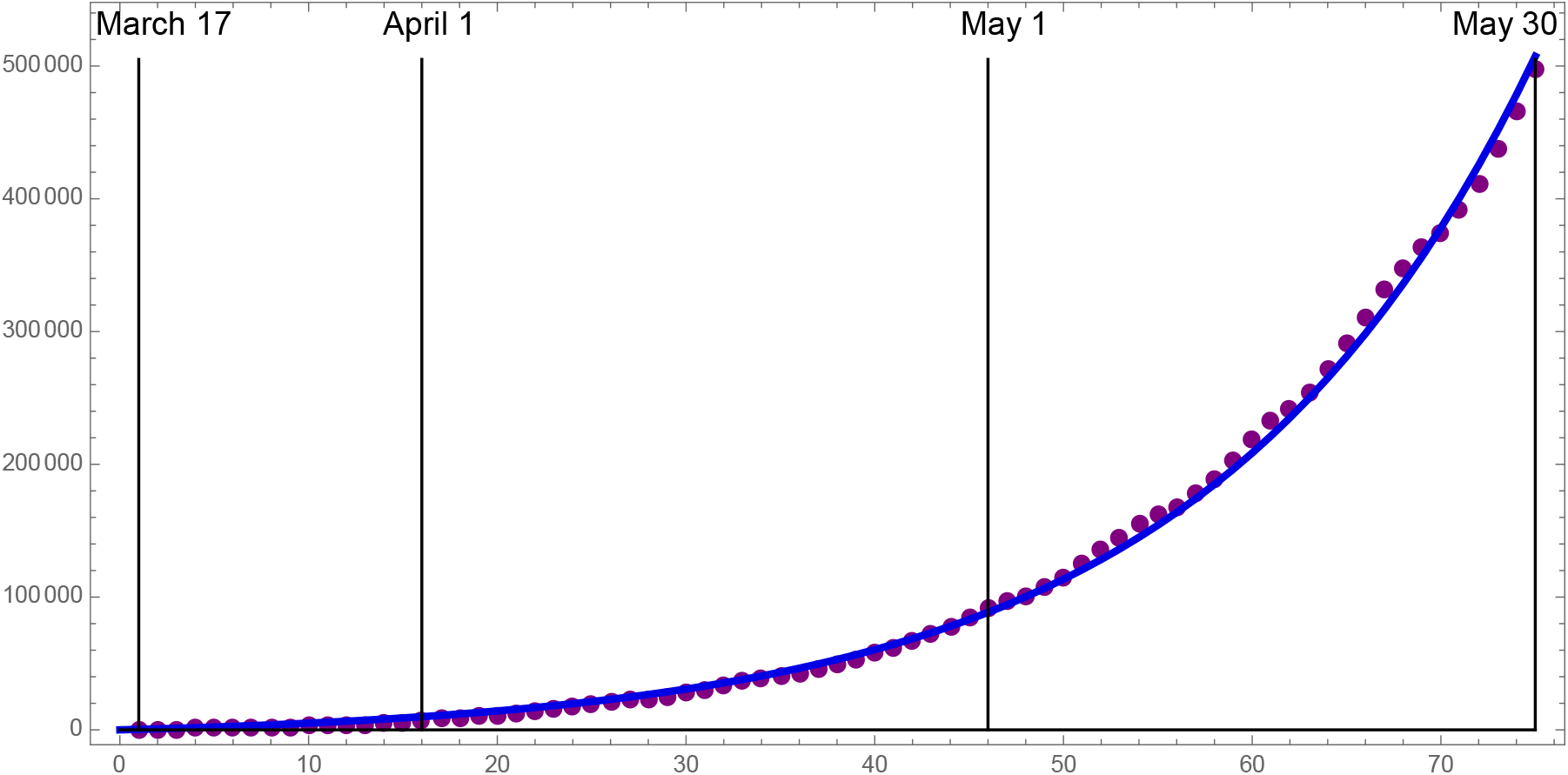
Dots: cumulative reported cases data from Tables 1,2,3. Graph: model simulation of the cumulative reported cases 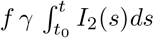, with *f* = 0.3 and γ =1/7. f is the fraction of removed *I*_2_ cases that are reported.

**Figure 3:**
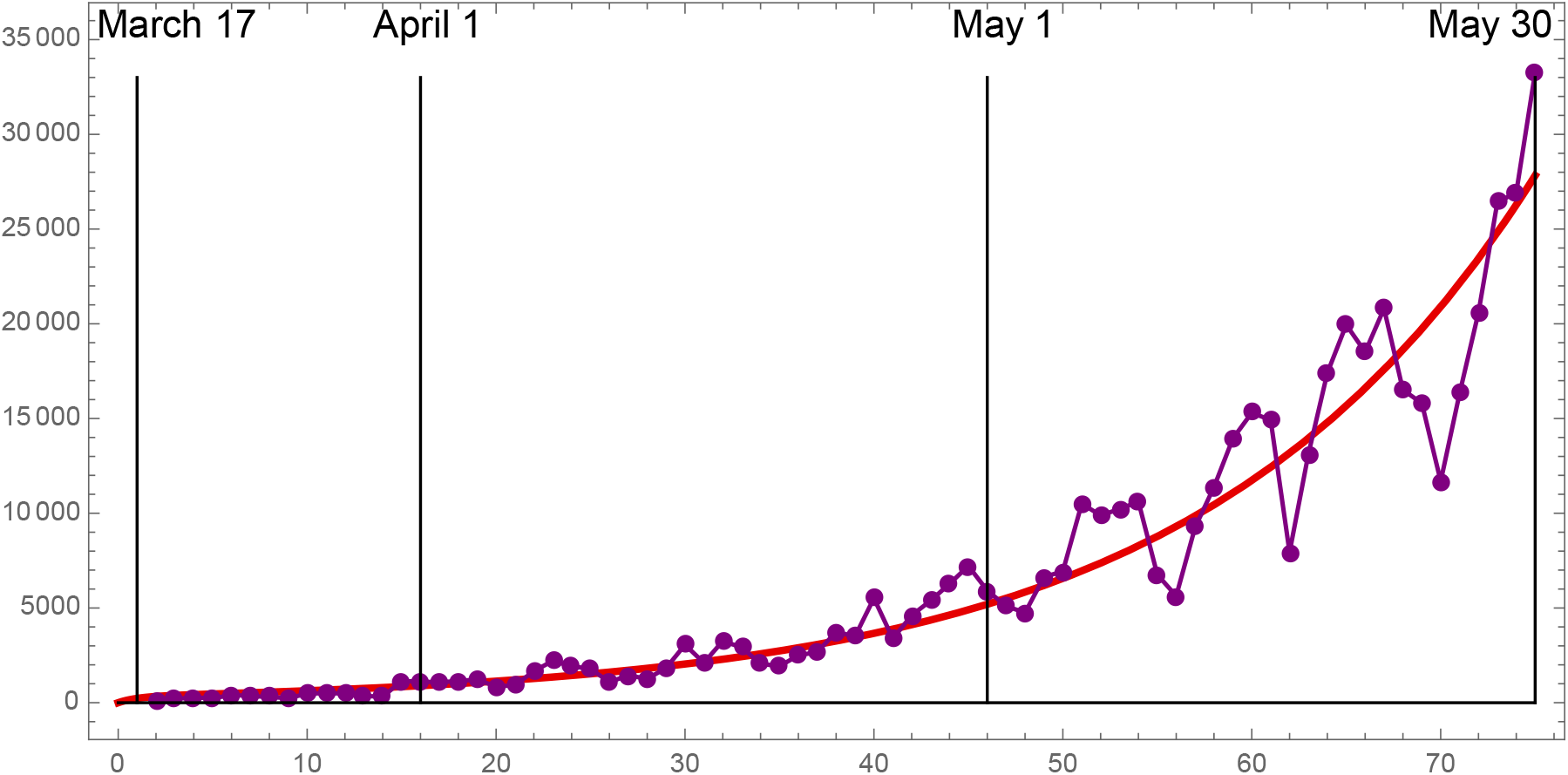
Dots: daily reported cases data obtained by subtraction day by day from the data in Tables 1,2,3. Graph: model simulation of the daily reported cases *DR*(*t*) with *f* = 0.3, γ = 1/7, and *t*_0_ = 1 (March 17).

**Table 1:** March - cumulative reported cases in Brazil.

|  |  |  |  |  |  |  |  |  |  |  |
| --- | --- | --- | --- | --- | --- | --- | --- | --- | --- | --- |
| 17 | 18 | 19 | 20 | 21 | 22 | 23 | 24 | 25 | 26 | 27 |
| 291 | 428 | 621 | 904 | 1128 | 1546 | 1891 | 2201 | 2433 | 2915 | 3417 |
| 28 | 29 | 30 | 31 |  |  |  |  |  |  |  |
| 3903 | 4256 | 4579 | 5717 |  |  |  |  |  |  |  |

**Table 2:** April - cumulative reported cases in Brazil.

|  |  |  |  |  |  |  |  |  |  |
| --- | --- | --- | --- | --- | --- | --- | --- | --- | --- |
| 1 | 2 | 3 | 4 | 5 | 6 | 7 | 8 | 9 | 10 |
| 6834 | 7910 | 9056 | 10278 | 11130 | 12056 | 13717 | 15927 | 17857 | 19638 |
| 11 | 12 | 13 | 14 | 15 | 16 | 17 | 18 | 19 | 20 |
| 20727 | 22169 | 23430 | 25262 | 28320 | 30425 | 33682 | 36599 | 38654 | 40581 |
| 21 | 22 | 23 | 24 | 25 | 26 | 27 | 28 | 29 | 30 |
| 43079 | 45757 | 49492 | 52995 | 58509 | 61888 | 66501 | 71886 | 78162 | 85380 |

**Table 3:**
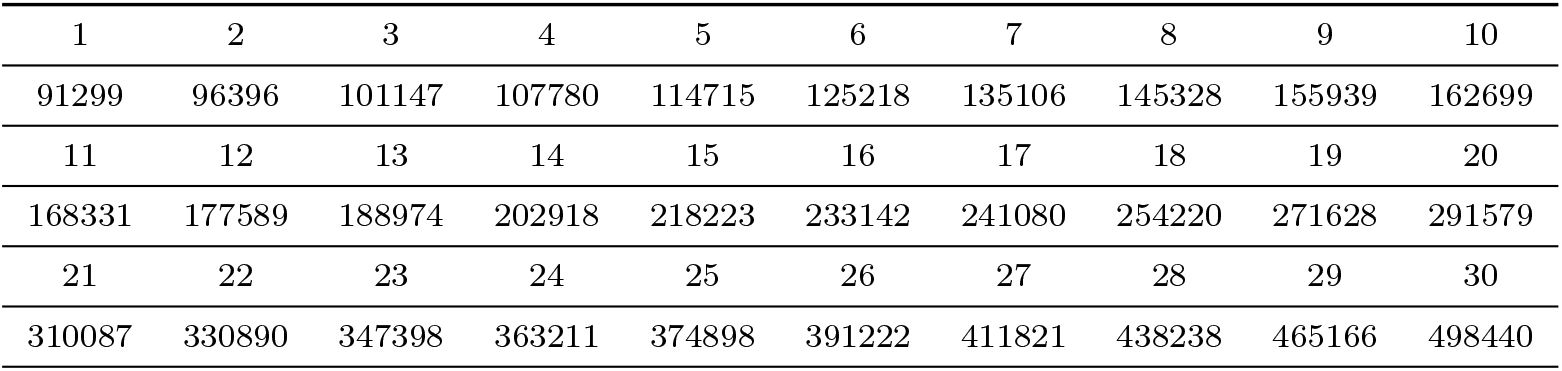
May - cumulative reported cases in Brazil.

In Figure 4 we project forward in time the model simulation of the cumulative reported cases 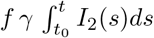, the model simulation of the total cumulative *I*_1_(*t*) cases 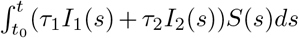, and the model simulation of the total *I*_2_(*t*) cases 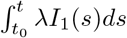. The final size of the epidemic is approximately 157,000,000 cases. The turning points of the cumulative cases are the times at which the graphs transition from concave up to concave down. The turning points are obtained by setting the second derivative of these graphs to 0. The turning points are graphed as vertical lines. The turning point of the cumulative *I*_2_ cases is approximately one week later than the turning point of the cumulative *I*_1_ cases, which is consistent with the assumption that the average time of *I*_1_ infectiousness 1/λ = 7 days. The turning point of the reported cumulative *I*_2_ cases is approximately one week later than the turning point of the cumulative *I*_2_ cases, which is consistent with the assumption that the average time of *I*_2_ infectiousness 1/γ = 7 days.

**Figure 4:**
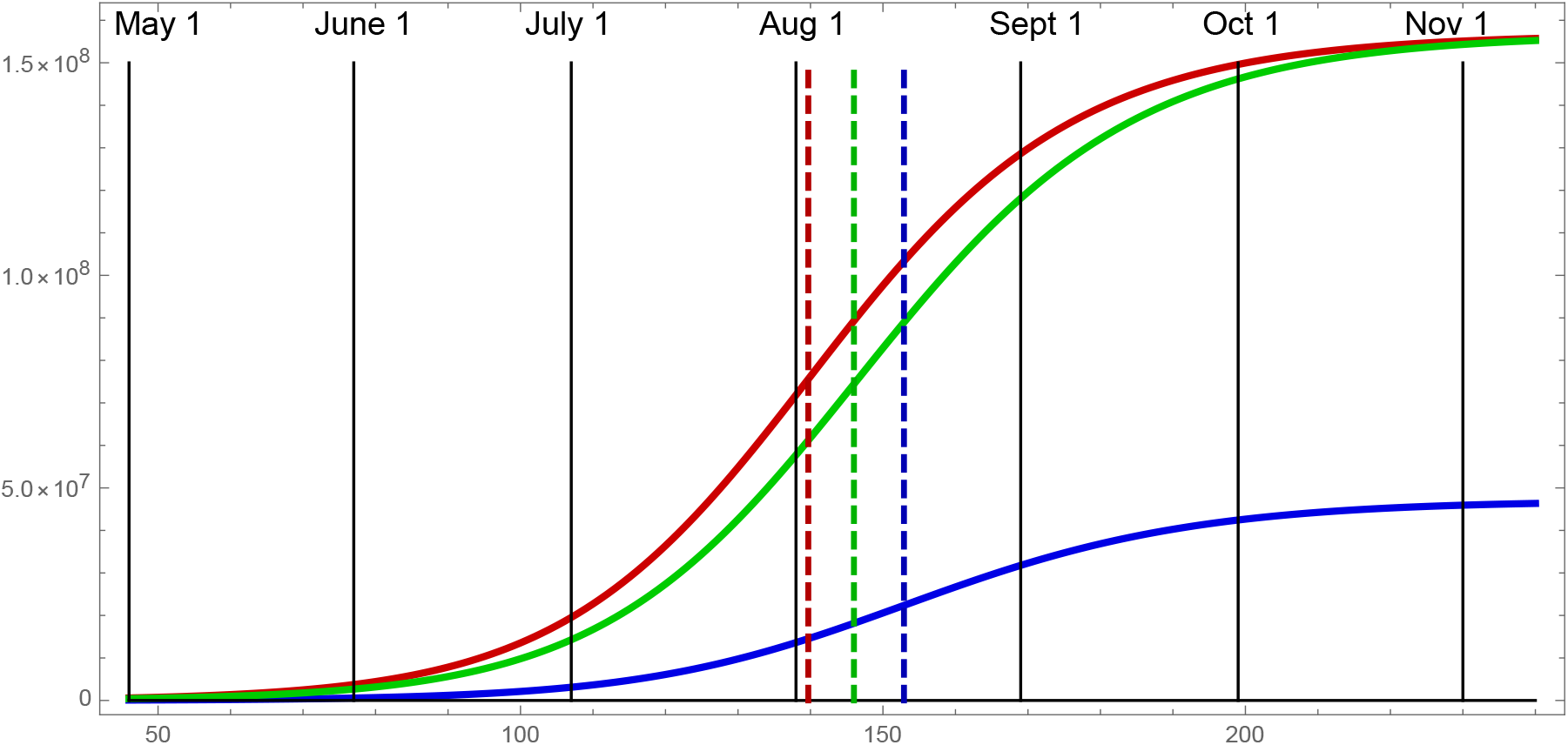
Blue: model simulation of the cumulative reported cases. Red: model simulation of the cumulative *I*_1_ infectious cases. Green: model simulation of the cumulative *I*_2_ infectious cases. The turning point of the cumulative reported cases is approximately day 152.6, the turning point of the cumulative *I*_1_ cases is approximately day 139.5, and the turning point of the cumulative *I*_2_ cases is day 146. The lim*_t_*_→∞_*I*_2_(*t*) is approximately 157,000,000 cases.

The extremely high number of cases projected in the simulation, with on-going unaltered transmission rates, is very unlikely. It is certain that government imposed and socially adopted distancing measures will take effect, and mitigate transmission. Such social distancing measure result in reduction of both *τ*_1_ and *τ*_2_. Measures such as isolation and contact tracing of high level symptomatic *I*_2_ cases, and quarantining of low level symptomatic *I*_1_ cases, can be quantified in terms of the final size formula (Corollary 1.2) of the epidemic. The reduction of *τ*_2_, corresponds to the identification and isolation of *I*_2_ high level symptomatic cases. The reduction of *τ*_1_, corresponds to the contact tracing of *I*_2_ high level symptomatic cases, and monitoring and quarantining of those contact traced *I*_1_ asymptomatic or low level symptomatic cases. The formula (1.2) for *S*_∞_ in Corollary 1.2 can be used to estimate the effects of these mitigations. The values of the initial conditions in formula (1.3) can be reset to up-dated values, with modified parameters *τ*_1_,*τ*_2_, to give time-forward predictions of the final size *S*_0_ − *S_∞_*. The spatially homogeneous model (1.1) can thus be used to predict the final size of the epidemic in Brazil, as new data becomes available, corresponding to social distancing changes in the population.

In Figure 5 we use formula (1.2) to graph the final size *S*_0_ − *S*_∞_ as a function of the *I*_1_ transmission rate *τ*_1_ and the *I*_2_ transmission rate *τ*_2_. We assume that the initial values *I*_1_(0) = 13,000 and *I*_2_(0) = 8,200, and the parameters λ =1/7 and γ =1/7 are at baseline values. If the transmission rate *τ*_1_ of low level infectious individuals *I*_1_ is reduced to 1.5 × 10^−10^, and the transmission rate *τ*_2_ of high level infectious individuals *I*_2_ is reduced to 0.6 × 10^−9^, then the final size *S*(0) − S_∞_ of the epidemic is approximately 38,000,000 cases. The effects of various transmission rate reductions can be viewed in the graph.

**Figure 5:**
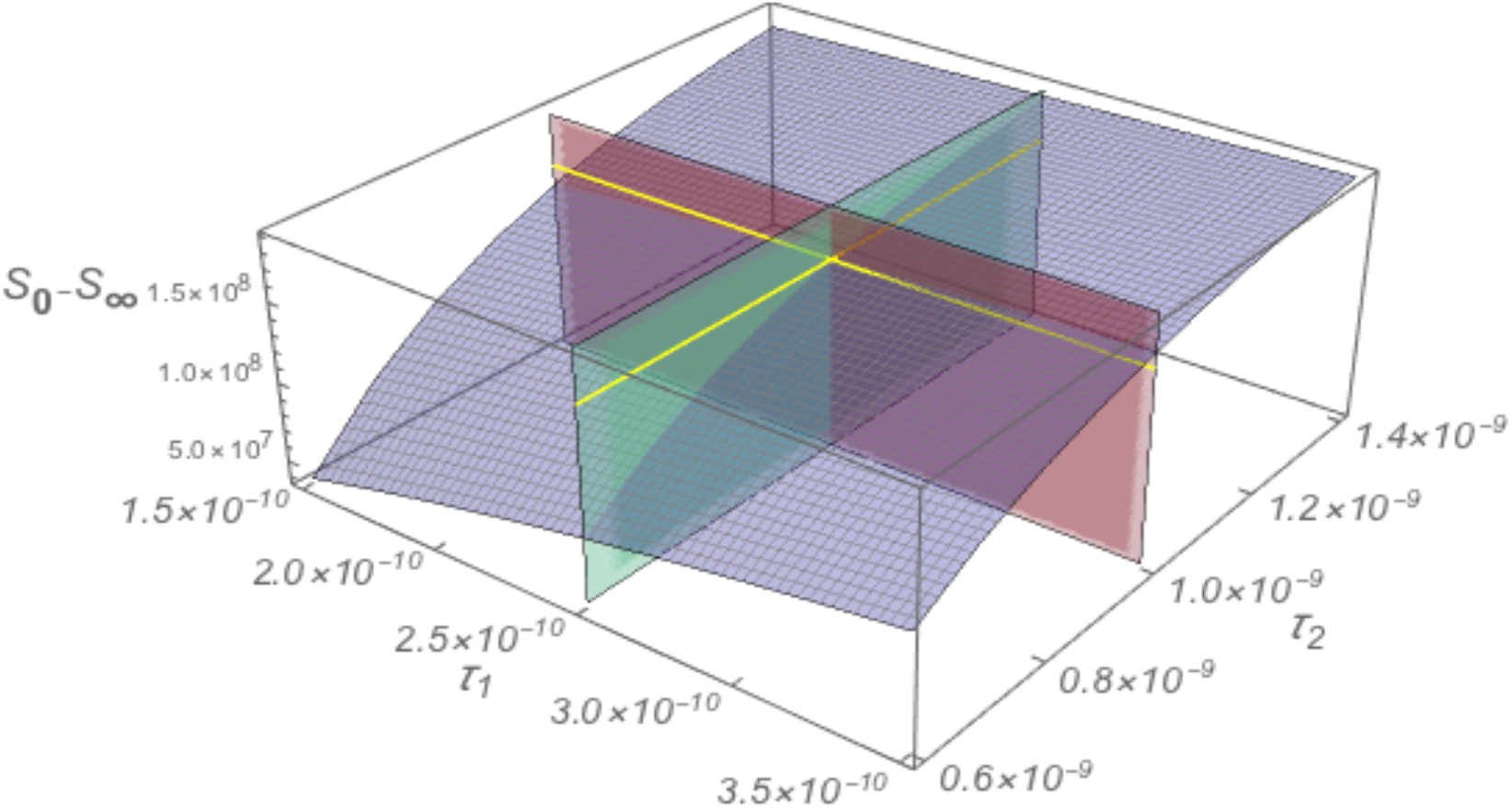
Blue surface: final size *S*_0_ − S_∞_ graphed as a function of of the transmission parameters *τ*_1_ and *τ*_2_. The yellow lines and red and green planes correspond to the baseline values *τ*_1_ = 2.5 × 10^−10^, *τ*_2_ = 1.0 × 10^−9^, *S*_0_ − S_∞_ = 157,000,000. The final size *S*(0) − S_∞_ decreases greatly as *τ*_1_ decreases and as *τ*_2_ decreases.

In Figure 6 we graph the final size *S*_0_ − S_∞_ as a function of the transmission parameters *τ*_1_ and *τ*_2_, with one fixed at baseline, and the other decreasing from baseline. We assume the initial values *I*_1_(0) = *I*_1_(*t*_1_) = 969,000 and *I*_2_(0) = *I*_2_(*t*_1_) = 692,000 in formula (1.3) are reset values at time *t*_1_ = May 30, in the baseline model simulation. We assume λ =1/7 and γ =1/7 are at baseline values. The reduction of the symptomatic class *I*_2_ transmission rate *τ*_2_ has greater effect in reducing the final size, than the reduction of the asymptomatic class *I_I_* transmission rate *τ*_1_, because prior to May 30, *I*_2_ transmissions were largely unrestrained by distancing measures.

**Figure 6:**
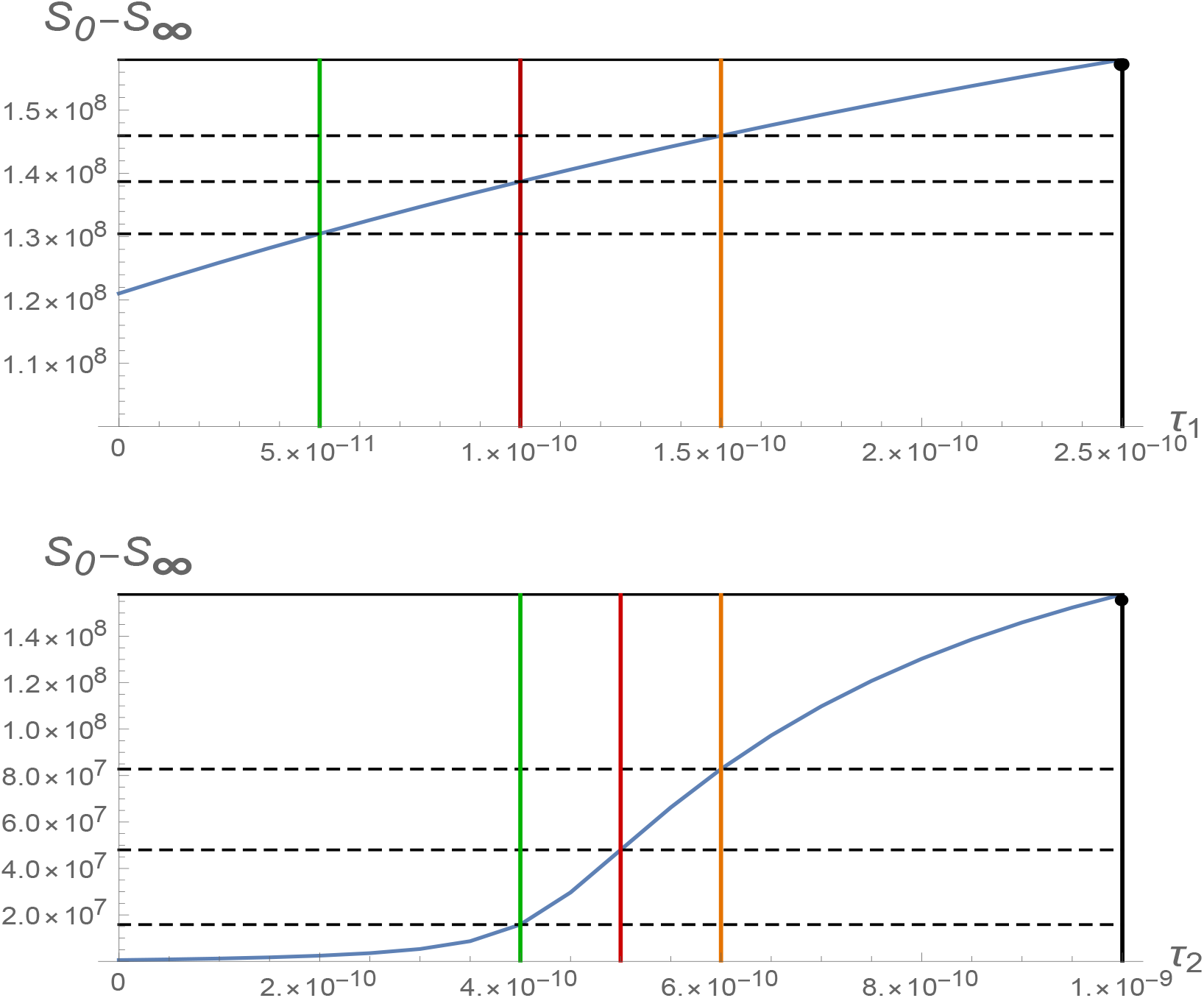
Blue surface: final size *S*_0_ − S_∞_ graphed as a function of *τ*_1_ and *τ*_2_. The yellow lines and red and green planes correspond to the baseline values *τ*_1_ = 2.5 × 10^−10^, *τ*_2_ = 1.0 × 10^−9^, *S*_0_ − S_∞_ = 157,000,000. The final size *S*(0) − *S*_∞_ decreases greatly as *τ*_1_ decreases and as *τ*_2_ decreases. For *τ*_1_ = 1.5 × 10^−10^ and *τ*_2_ = 0.6 × 10^−9^, the final size *S*(0) − S_∞_ is approximately 38, 000,000.

In Figure 7 we graph the basic reproductive number 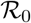 in formula (1.3) as a function of the transmission parameters *τ*_1_ and *τ*_2_. 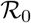 is, approximately, the number of transmissions generated by one infectious individual in the outbreak stage of the epidemic. The reduction of the symptomatic class *I*_2_ transmission rate *τ*_2_ has greater effect in reducing 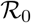, than the reduction of the asymptomatic class *I_I_* transmission rate *τ*_1_, because prior to May 30, *I*_2_ transmissions were largely unrestrained by distancing measures.

**Figure 7:**
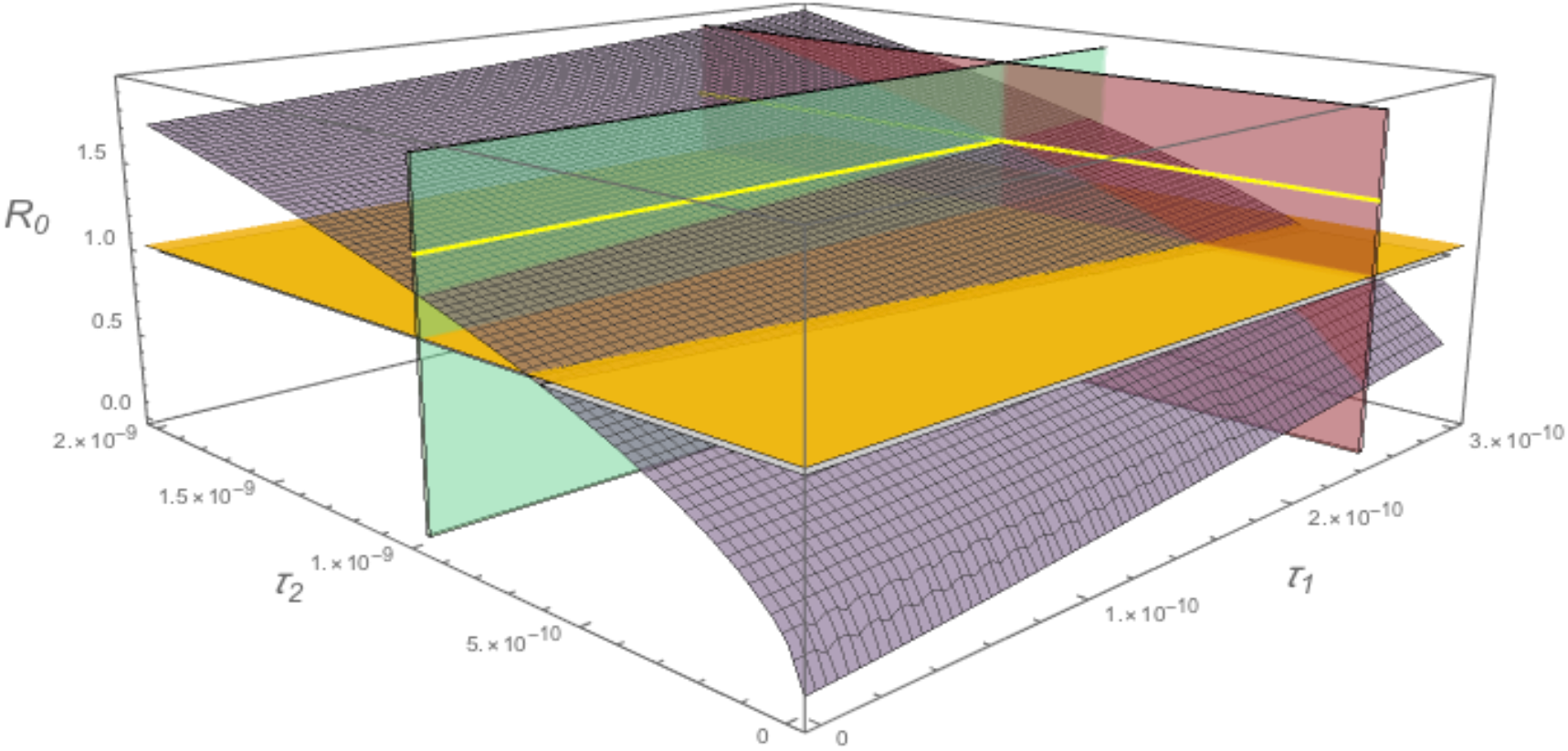
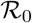 as a function of the transmission parameters *τ*_1_ and *τ*_2_. The yellow lines and red and green planes correspond to the baseline values *τ*_1_ = 2.5 × 10^−10^, *τ*_2_ = 1.0 × 10^−9^. The orange plane is at 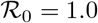. At baseline, 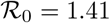. The epidemic transmission is mitigated if 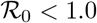.

## 3 Conclusions

We have developed a dynamic model of a COVID-19 epidemic outbreak in a susceptible population. Our model describes the outbreak as a growth of transmission of susceptible individuals from two classes of infectious individuals: *I*_1_ asymptomatic (low level symptomatic), and *I*_2_ symptomatic (high level symptomatic). The model does not assume that major government measures or social behaviour changes have been implemented to mitigate the epidemic transmission. The model also assumes that reported cases are a fraction of the total cases in the population.

We apply the model to the COVID-19 current in Brazil. We identify parameters and initial conditions that give agreement of the model output with current case data from Ministerio da Saude of Brazil. We project forward in time the model solutions, to give final size predictions of the epidemic in the absence of social distancing measures. The extreme scenario that the epidemic continues, without social distancing mitigation, is extremely unlikely, given the magnitude of this final size. Intervention measures, such as isolation and contact tracing of high level symptomatic *I*_2_ cases, and quarantining of low level symptomatic *I*_1_ cases, can be quantified in terms of the final size formula (Corollary 1.2) of the epidemic. The reduction of *τ*_2_, corresponds to the identification and isolation of *I*_2_ high level symptomatic cases. The reduction of *τ*_1_, corresponds to the contact tracing of *I*_2_ high level symptomatic cases and quarantining of contact traced *I*_1_ asymptomatic or low level symptomatic cases. Both measures have major effect in controlling the epidemic.

## Data Availability

All data in the manuscript is available upon request to the corresponding author.

https://coronavirus.saude.gov.br/

## Notes

### Competing Interest Statement

The authors have declared no competing interest.

### Clinical Trial

No clinical trials were performed.

### Funding Statement

No external funding was received.

### Author Declarations

All relevant ethical guidelines have been followed.

